# The uncertain self: intolerance of uncertainty moderates the association of positive schizotypal traits and self-concept clarity

**DOI:** 10.1101/2025.08.01.25332788

**Authors:** Alexandra Kaszás, Szabolcs Kéri, Dorottya Bencze, Ágnes Szőllősi, Ágota Vass, Mihály Racsmány, Bertalan Polner

## Abstract

Our objective was to investigate the relationship between self-concept clarity, intolerance of uncertainty, and schizotypal traits, with a focus on whether intolerance of uncertainty moderates the association between self-concept clarity and positive schizotypy in a non-clinical context. A sample of 316 adults (on average 43 (SD=12) years, 247 women) completed the Self-Concept Clarity Scale, the Intolerance of Uncertainty scale, and the Schizotypal Personality Questionnaire - Brief Revised. Lower self-concept clarity was significantly associated with higher levels of positive, negative, and disorganized schizotypy, as well as with greater inhibitory intolerance of uncertainty. Importantly, a moderation analysis revealed that inhibitory intolerance of uncertainty significantly altered the strength of the negative relationship between self-concept clarity and positive schizotypy. Specifically, this association was strongest among individuals with low levels of inhibitory intolerance of uncertainty. In contrast, at higher levels of inhibitory intolerance of uncertainty, the negative relationship between self-concept clarity and positive schizotypy was weaker. This pattern suggests that among those who are less behaviorally inhibited in face of uncertainty, a better defined self-concept is more strongly coupled with lower levels of positive schizotypy. These results underscore the importance of considering both self-structure and uncertainty tolerance in models of schizotypy and psychosis risk. Our findings suggest that self-concept clarity and intolerance of uncertainty are interrelated cognitive factors that synergistically explain variance in schizotypal traits. As intolerance of uncertainty is a potentially modifiable cognitive process, the results have implications for early interventions aimed at reducing psychosis risk by improving self-concept clarity and tolerance for uncertainty. Although the study used a non-clinical sample, it supports a dimensional approach to psychosis and highlights key targets for future interventional research.

## Introduction

The concept of subjectivity—the lived experience of being a self-aware, thinking, feeling individual— and its disorders have become a prominent topic in psychological and psychiatric research, gaining increasing attention from both researchers and the general public. However, schizophrenia—a disorder of subjectivity in which the connection between an individual’s inner experiences and the external world is altered, leading to a distorted perception and interpretation of reality—remains one of the most misunderstood and stigmatized mental illnesses, leaving many open questions for researchers. Studying patients diagnosed with schizophrenia presents significant challenges due to the severity of the illness, poor patient conditions, potential substance abuse, hospitalization, and the side effects of medication. [1]

To address these challenges, researchers have developed various animal and human models to investigate psychotic mechanisms. These models fall into two categories: state models, which examine transient conditions, and trait models, which study persistent traits such as high levels of schizotypy in the general population [2]. Schizotypy exists on a continuum between normality and schizophrenia [3]. This dimensional perspective allows researchers to investigate psychosis-proneness without the confounding effects of chronic illness, medication, or hospitalization commonly present in clinical samples. Studying schizotypal traits in non-clinical populations thus provides a unique opportunity to explore the psychological and cognitive mechanisms underlying psychosis risk in a more controlled and generalizable context. Moreover, it facilitates early identification of vulnerability markers and contributes to our understanding of how certain personality traits may evolve into more severe psychopathological conditions [1].

Like schizophrenia, schizotypy can be divided into three dimensions based on factor analysis: positive, negative, and disorganized schizotypy [4]. Positive schizotypy involves an excess of phenomena that should not be present, such as unusual experiences, magical thinking, or ideas of reference [5]. In contrast, negative schizotypy reflects deficits in expected functions, such as avolition or an absence of close relationships [6]. Disorganized schizotypy pertains to disturbances in the regulation of thought and behavior [7]. Notably, higher levels of disorganization may predict increased positive and negative traits, and vice versa [6]. As a result, disorganization must be considered to fully differentiate the dimensions of schizotypy.

A central topic in this field is the concept of the self, as schizophrenia is often described as a disorder of self-fragmentation [8]. The self can be defined as “the totality of the individual, consisting of all characteristic attributes, conscious and unconscious, mental and physical”. However, interpretations of the self vary among theorists. Heinz Kohut, for instance, defined it as a “coherent, stable, yet dynamic experience of one’s individuality, continuity in time and space, autonomy, efficacy, motivation, values, and desires” [9]. Similarly, Bleuler [8,10] suggested that disruptions in personality unity may lead to a loss of self-concept clarity.

Self-concept clarity (SCC) is a central structural feature of the self, defined as the extent to which an individual’s beliefs about themselves are clearly and confidently defined, internally consistent, and stable over time [11]. A healthy self-concept allows an individual to perceive themselves as a coherent and consistent being, maintain a stable yet flexible identity, distinguish themselves from others, and regulate thoughts, emotions, and actions. Consistent with this, SCC has been shown to correlate with a broad range of psychological outcomes, including resilience, well-being, interpersonal functioning, and emotional adjustment [12,13].

The link between schizophrenia and self-concept disruption has been recognized since the 1950s [12,14]. One of the most influential studies on this topic, conducted by Cicero et al. [15], found that while evaluative components of self-concept (e.g., self-esteem, self-congruence) do not distinguish individuals with schizophrenia from healthy controls, structural components (e.g., organization) play a crucial role in symptom development, particularly in non-chronic schizophrenia. Several studies have confirmed that individuals on the schizophrenia spectrum exhibit lower self-concept clarity during their first psychotic episode [15,16]. While some researchers found that low self-concept clarity correlates with increased psychotic symptoms [16], others reported that self-concept clarity was associated with increased positive symptoms and a higher subjective quality of life—but only under low stress [17]. These findings suggest that the relationship between self-concept clarity and positive symptoms is more complex than initially thought, warranting further investigation.

In stressful situations, individuals with schizophrenia exhibit a less stable self-concept, making them more vulnerable to the negative effects of failure. This instability is not merely a general response but a specific cognitive vulnerability within schizophrenia [18]. When self-concept is unstable, individuals become more prone to anxiety and struggle with future planning [12]. In this context, self-concept clarity can be understood as the degree to which one is certain about their sense of self.

Low self-concept clarity may impair an individual’s ability to navigate the world with a stable identity, increasing their anxiety in uncertain situations [19]. This anxiety can contribute to intolerance of uncertainty, which refers to how individuals perceive and react to ambiguous or unpredictable situations [20]. Those with high intolerance of uncertainty often find unpredictable events distressing and upsetting. Research suggests that this tendency is more strongly linked to intolerance of uncertainty than to general anxiety or worry [21]. Studies on clinical populations indicate that the severity of delusional thinking is associated with higher levels of intolerance of uncertainty [22,23]. Additionally, White and Gumley [24] found that distress associated with psychosis was linked to intolerance of uncertainty, which, in turn, correlated with a perceived loss of control. This may explain why rigid or delusional beliefs can provide temporary relief by creating a sense of certainty [18].

Since intolerance of uncertainty has been linked to positive symptoms of schizophrenia, an important question arises: could another key feature—self-concept clarity—also be related? However, only two studies have directly examined the relationship between self-concept clarity and intolerance of uncertainty [255,26]. Butzer and Kuiper [26], in a study with 166 participants, found a negative association between these two factors in the general population, while both of them were related to anxiety and depression. Kusec et al. [25] further explored this connection in the context of generalized anxiety disorder (GAD). Their findings showed that individuals with high GAD symptoms exhibited two distinguishing traits: increased intolerance of uncertainty and reduced self-concept clarity.

These studies provide theoretical grounding for why intolerance of uncertainty may be a critical moderator. When individuals encounter events that contradict their expectations, they often respond by engaging in various strategies to restore a sense of coherence or stability. People with high intolerance of uncertainty are more sensitive to violations of internal coherence (like an unclear self-concept), which could amplify unusual schizotypal experiences and magical thinking [27].

Given the limited research on this topic, we aim to expand the literature by examining the relationship between self-concept clarity and intolerance of uncertainty within the context of schizotypy. We used validated scales and questionnaires in a sample of 316 participants, to investigate whether intolerance of uncertainty moderates the relationship between self-concept clarity and positive schizotypy. Based on the aforementioned literature and a review of the results, we hypothesize that disturbances in self-concept clarity are negatively associated with both schizotypy and intolerance of uncertainty. Furthermore, we expect that intolerance of uncertainty moderates the relationship between self-concept clarity and positive schizotypy in that the association will be particularly strong in those with low intolerance of uncertainty.

## Method

### Participants

Participants aged 18 and over were recruited between September 4 and December 31, 2022, through advertisements posted on the research team’s Facebook page. Here, we analyse data from a study originally designed to study the associations between schizotypal traits and memory performance (will be reported in a separate study). In the context of that research question, based on a power analysis (the univariate association between schizotypy and visual memory alterations is expected to be “small”: r = 0.2, alpha = 0.05, beta = 0.1 [28]), the minimum required sample size was determined to be 259 participants. Because the association between schizotypy and other self-reported traits is expected to be in general higher than that of schizotypy and memory due to common method variance, the above estimation of 90% (1 - beta * 100) provides a lower bound for the statistical power obtained for the bivariate associations in this study. Recruitment was carried out using a convenience sampling method, with participants recruited through personal networks and paid advertisements. No restrictions were applied regarding participants’ personal interests or demographic characteristics. Participation was voluntary, and anonymity was ensured. Before beginning the survey, participants received detailed information about the study. They were required to explicitly indicate their informed consent by checking boxes confirming that they had read the information and agreed to participate. A password system was implemented to prevent duplicate entries. The study adhered to the ethical principles outlined in the Helsinki Declaration and was approved by the United Ethical Review Committee for Research in Psychology, Hungary (2016/032, 2019/29).

Initially, 317 participants completed the study; however, one participant was excluded due to providing identical responses to all items on the Self-Concept Clarity Scale, including reversed items. As a result, the final sample consisted of 316 participants (self-reported sex: 68 men, 247 women, and 1 non-binary). The average age of the participants was 43 years, with a range from 18 to 74 years (SD = 11.8 years). In terms of education, one-third (32%) of the participants held a Bachelor’s degree, another third (32%) had a higher education degree, and the remaining third (36%) had lower education. Twenty-eight percent of the respondents reported being currently enrolled in educational studies. Geographically, 32% of the sample resided in the capital, 54% lived in other cities or towns, and the remaining participants (14%) lived in settlements with fewer than 1,000 inhabitants. In terms of marital status, 59% of participants were married or in a relationship, 28% were single, and 13% were divorced or widowed.

### Questionnaires

Schizotypy was assessed using the Schizotypal Personality Questionnaire - Brief Revised (SPQ-BR; [29,30]). The 32 items of the questionnaire can be grouped in two ways: higher-order and lower-order subscales. The higher-order subscales measure positive (14 items), negative/interpersonal (10 items), and disorganized (8 items) schizotypy, while the lower-order subscales include ideas of reference, suspiciousness, magical thinking, unusual perceptions, social anxiety, constricted affect/lack of close friends, eccentric behavior, and odd speech. As the lower-order subscales consist of only a few items, we followed the tradition in the literature and used only the higher-order subscales (*SPQ positive: α = 0.75, 95% CI [0.70– 0.79]; SPQ negative: α = 0.76, 95% CI [0.72–0.80]; SPQ disorganized: α = 0.71, 95% CI [0.66–0.76]*). Participants were asked to rate their agreement with each statement on a 5-point Likert scale ranging from 1 to 5.

Self-concept clarity was measured using the Self-Concept Clarity Scale (SCCS; [11,31]), which assesses the clarity and strength of individuals’ beliefs about themselves using 12 items (*α = 0.90, 95% CI [0.88–0.91]*). Participants rated each item on a scale from 1 (=disagree) to 5 (=agree).

Intolerance of uncertainty was assessed using the short version of the Intolerance of Uncertainty Scale (IUS-12; [32,33]), which measures individuals’ ability to tolerate the occurrence and unpredictability of negative events. The 12 items are divided into two subscales: prospective and inhibitory anxiety. The prospective subscale (IUS PRO) includes 7 items about the fear of future events, while the inhibitory subscale (IUS INH) of 5 items inquires about questions about the incapacity to act caused by uncertainty (*IUS PRO: α = 0.72, 95% CI [0.66–0.78]; IUS INH: α = 0.78, 95% CI [0.73–0.82]*). Participants rated each statement on a scale from 1 (=strongly disagree) to 5 (=strongly agree), depending on the statement they agreed with the most. Based on Cronbach’s α values, all main subscales met the 0.7 threshold, indicating good internal consistency and reliability.

### Procedure

The test materials were administered to participants using the open-source software *formr* [34], where the data were also stored. This platform provides researchers with an innovative tool for designing questionnaires and tests, distributing them to participants, and enhancing reproducibility. We utilized the software to monitor data collection and track participant feedback. Questionnaires were administered in a fixed order: SCCS, IUS-12, and SPQ-BR.

### Statistical analysis

Our research questions were tested using data analysis in JASP (version 0.19.3.0) [35], and data visualization was conducted using the R programming language. The Shapiro-Wilk test indicated that normality was violated for all investigated variables, which was accounted for in the analysis. Additionally, variable distributions were assessed visually.

The relationships between schizotypy dimensions and self-concept clarity, as well as between self-concept clarity and intolerance of uncertainty, were examined using the robust Spearman’s correlation. The potential moderating role of intolerance of uncertainty in the relationship between the positive dimension of schizotypy and self-concept clarity was assessed through linear regression models. These models were adjusted for age and sex, as previous research indicates associations between these factors and schizotypy [36] as well as self-concept clarity [37,38]. A 95% confidence interval was calculated.

## Results

We examined the relationship between self-concept clarity and the three dimensions of schizotypy using Spearman’s correlation with partialization. Confidence intervals were estimated based on 5000 bootstrap samples. This analysis aimed to establish a foundation for our additional research questions. To account for potential confounders, we controlled for sex and age. As shown in Fig 1 and Table 1, the results indicated that self-concept clarity was negatively associated with all three schizotypy dimensions: **positive** (*ρ = −0.529, p < 0.001, 95% CI [−0.614, −0.433]*), **negative** (*ρ = −0.506, p < 0.001, 95% CI [−0.594, −0.411]*), and **disorganized** (*ρ = −0.528, p < 0.001, 95% CI [−0.608, −0.436]*). All associations were strongly negative, suggesting that higher levels of schizotypy are associated with lower self-concept clarity, and vice versa.

**Fig 1.**
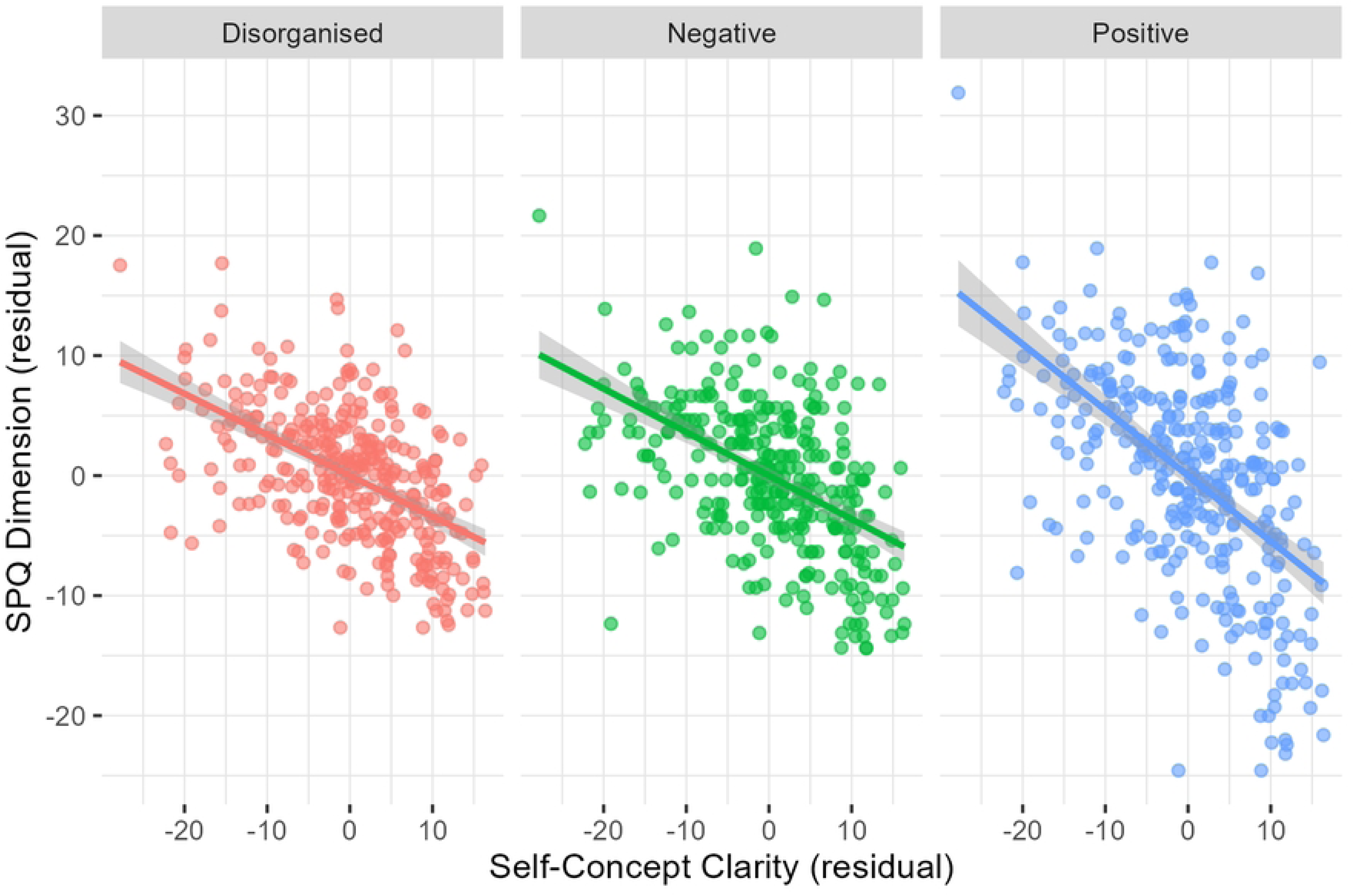
Partial correlations between SCCS and SPQ dimensions. *Note.* SCCS = Self-Concept Clarity Scale. SPQ = Schizotypy Personality Questionnaire. Scatter plots of residuals from regressions controlling for age and gender, showing partial correlations between SCCS and SPQ dimensions. Trend lines represent linear fits with 95% confidence intervals.

**Table 1.**
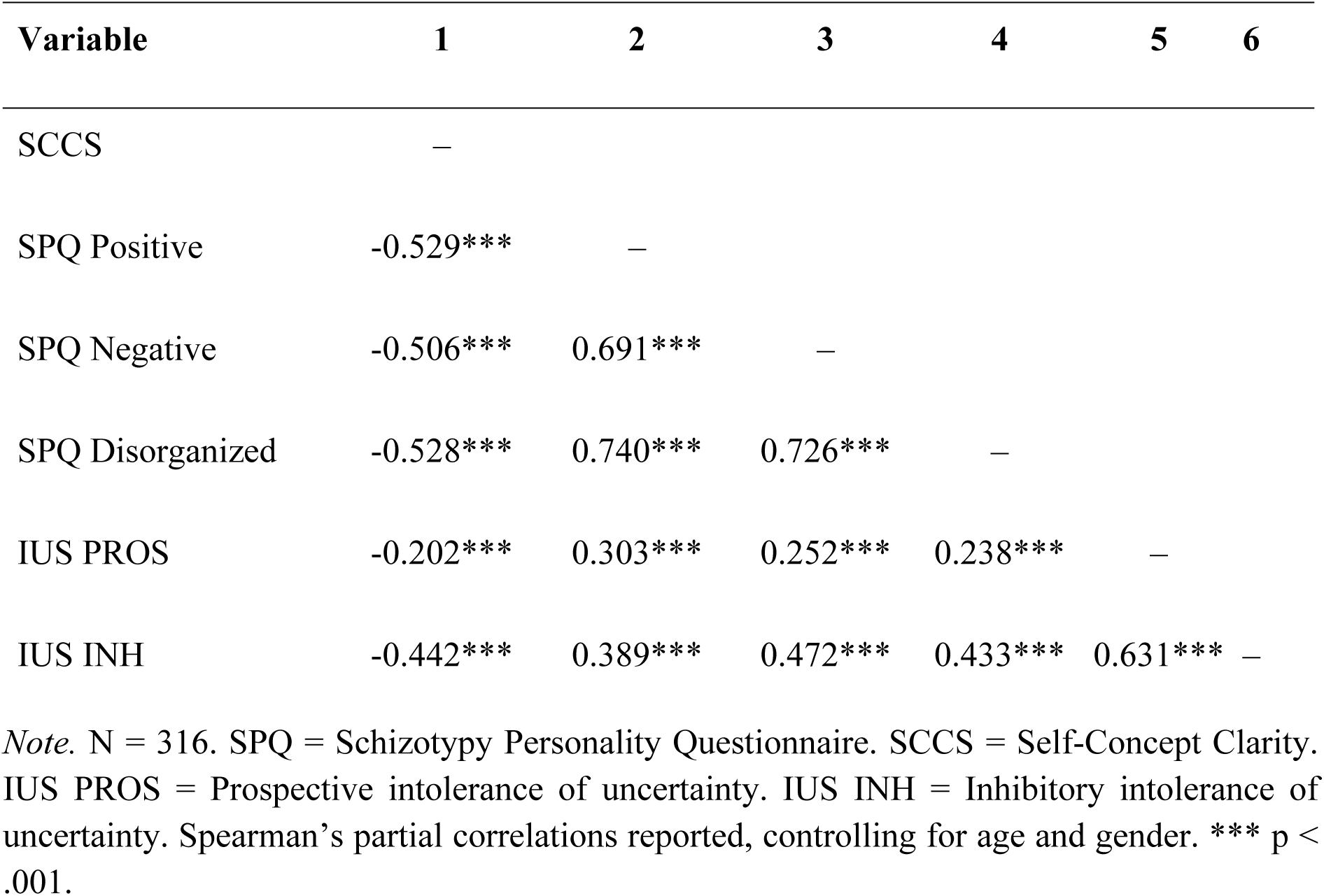
Spearman correlations between self-concept clarity, SPQ dimensions, and intolerance of uncertainty.

| Variable | 1 | 2 | 3 | 4 | 5 | 6 |
| --- | --- | --- | --- | --- | --- | --- |
| SCCS | — |  |  |  |  |  |
| SPQ Positive | -0.529*** | — |  |  |  |  |
| SPQ Negative | -0.506*** | 0.691*** | — |  |  |  |
| SPQ Disorganized | -0.528*** | 0.740*** | 0.726*** | — |  |  |
| IUS PROS | -0.202*** | 0.303*** | 0.252*** | 0.238*** | — |  |
| IUS INH | -0.442*** | 0.389*** | 0.472*** | 0.433*** | 0.631*** | — |

*Note.* N = 316. SPQ = Schizotypy Personality Questionnaire. SCCS = Self-Concept Clarity. IUS PROS = Prospective intolerance of uncertainty. IUS INH = Inhibitory intolerance of uncertainty. Spearman's partial correlations reported, controlling for age and gender. \*\*\* $p < .001$ .

Next, we examined the relationship between self-concept clarity and the two dimensions of intolerance of uncertainty using Spearman’s correlation, controlling for sex and age. Self-concept clarity exhibited a weak negative association with **prospective intolerance of uncertainty** (*ρ = −0.202, p < 0.001, 95% CI [−0.311, −0.090]*) and a moderate negative association with **inhibitory intolerance of uncertainty** (*ρ = −0.442, p < 0.001, 95% CI [−0.526, −0.344]*). The results are shown in Table 1 and Fig 2.

**Fig 2.**
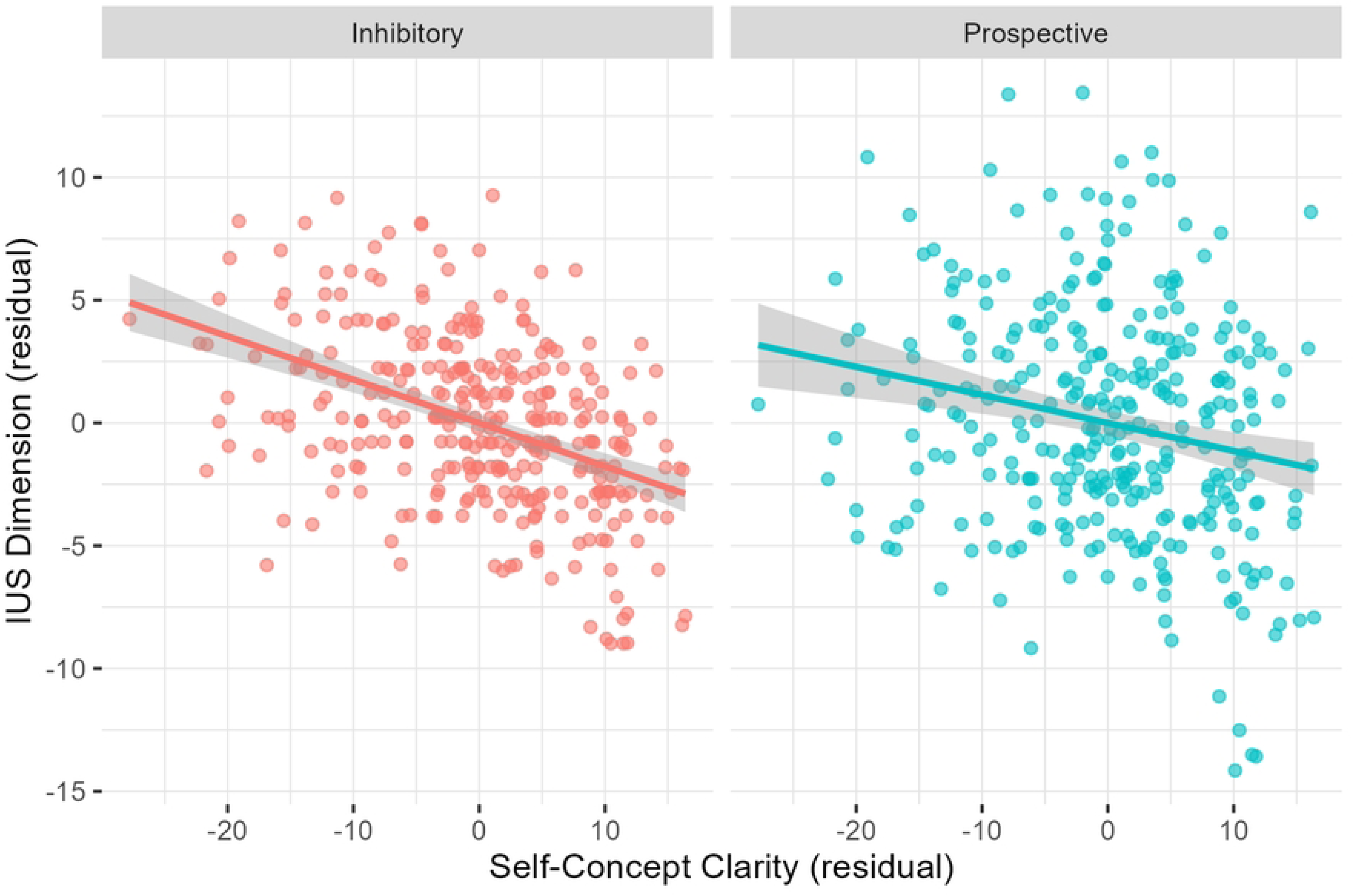
Partial correlations between SCCS and IUS dimensions. Note. SCCS = Self-Concept Clarity Scale. IUS = Intolerance of Uncertainty Scale. Scatter plots of residuals from regressions controlling for age and gender, showing partial correlations between SCCS and IUS dimensions. Trend lines represent linear fits with 95% confidence intervals.

To assess moderation effects, we conducted two linear regression analyses with the positive dimension of schizotypy as the dependent variable. Self-concept clarity and the two dimensions of intolerance of uncertainty were included as predictors in separate models: the first model incorporated the prospective dimension, while the second model included the inhibitory dimension. An interaction term was included to evaluate the moderation effect. Age, sex and the disorganized dimension were also controlled for to account for their potential influence.

The first model yielded the following values: R = 0.803; R² = 0.645; Adjusted R² = 0.638; RMSE = 5.347; *F*(X, N–X–1) = 93.501, *p* < 0.001. The second model produced similar values: R = 0.796; R² = 0.634; Adjusted R² = 0.627; RMSE = 5.428; *F*(X, N–X–1) = 89.210, *p* < 0.001. The R values indicate a strong relationship between the predictors and the positive dimension of schizotypy in both models. Collectively, the models explain approximately 63-64% of the variance in positive schizotypy, which is a substantial proportion. Additionally, the close alignment of the adjusted R² values with R² suggests that the predictors contribute meaningfully to explaining positive schizotypy and that the models are not overfitted.

The models are shown in Table 2A and 2B. In the first model, there was no significant interaction between self-concept clarity and prospective intolerance of uncertainty (*Beta = 0.014, SE = 0.007, Standardized Beta = 0.408, t = 1.843, p = 0.066*). In the second model, a significant interaction was observed between self-concept clarity and inhibitory intolerance of uncertainty. The interaction term yielded *Beta = 0.020, SE = 0.010, Standardized Beta = 0.352, t = 2.064, p = 0.040*. These findings suggest that, contrary to our hypothesis, higher levels of inhibitory intolerance of uncertainty dampen the association of self-concept clarity with positive schizotypy. Conversely, when intolerance of uncertainty is lower, the association of self-concept clarity with positive schizotypy is stronger. Fig 3 illustrates how different levels of inhibitory intolerance of uncertainty moderate this relationship.

**Table 2A.** Linear regression results for schizotypy with prospective IU as moderator.

| Predictor | B | SE B | $\beta$ | t | p |
| --- | --- | --- | --- | --- | --- |
| Intercept | 26.480 | 8.113 |  | 3.264 | 0.001 |
| SCCS | -0.464 | 0.166 | -0.477 | -2.803 | 0.005 |
| IUS PROS | -0.356 | 0.343 | -0.188 | -1.036 | 0.301 |
| Disorganized | 0.999 | 0.066 | 0.635 | 15.162 | < .001 |
| Age | 0.46 | 0.027 | 0.062 | 1.703 | 0.090 |
| Sex | -0.783 | 0.733 | -0.037 | -1.068 | 0.286 |
| SCCS x IUS PROS | 0.014 | 0.007 | 0.408 | 1.843 | 0.066 |

*Note.* N = 316. SCCS = Self-Concept Clarity. IUS PRO = Prospective Intolerance of Uncertainty. Disorganized = Disorganized dimension of schizotypy. B = unstandardized coefficients; SE B = standard error of B; $\beta$ = standardized coefficient.

**Table 2B.**
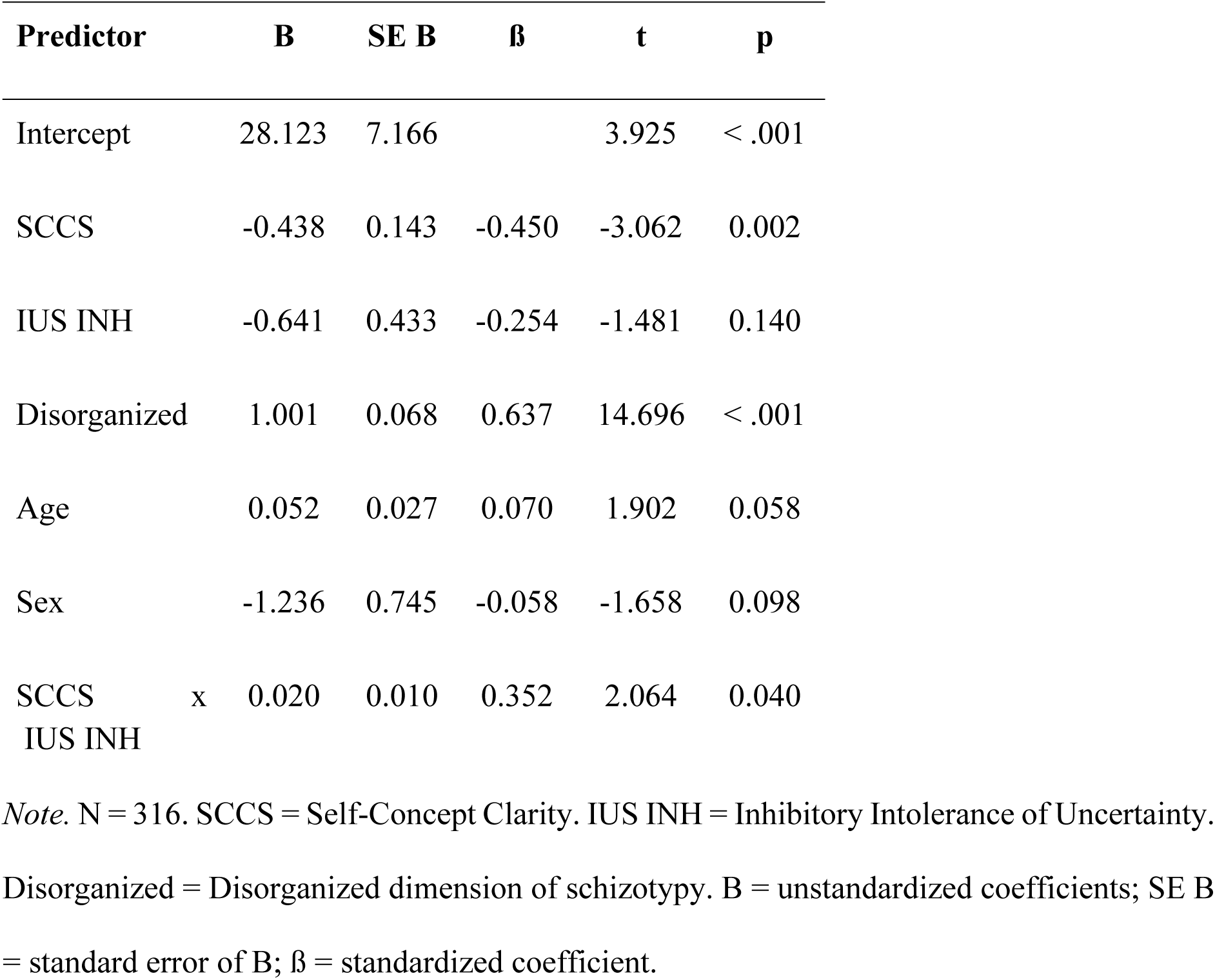
Linear regression results for schizotypy with inhibitory IU as moderator.

| Predictor | B | SE B | $\beta$ | t | p |
| --- | --- | --- | --- | --- | --- |
| Intercept | 28.123 | 7.166 |  | 3.925 | < .001 |
| SCCS | -0.438 | 0.143 | -0.450 | -3.062 | 0.002 |
| IUS INH | -0.641 | 0.433 | -0.254 | -1.481 | 0.140 |
| Disorganized | 1.001 | 0.068 | 0.637 | 14.696 | < .001 |
| Age | 0.052 | 0.027 | 0.070 | 1.902 | 0.058 |
| Sex | -1.236 | 0.745 | -0.058 | -1.658 | 0.098 |
| SCCS x IUS INH | 0.020 | 0.010 | 0.352 | 2.064 | 0.040 |

*Note.* N = 316. SCCS = Self-Concept Clarity. IUS INH = Inhibitory Intolerance of Uncertainty. Disorganized = Disorganized dimension of schizotypy. B = unstandardized coefficients; SE B = standard error of B; $\beta$ = standardized coefficient.

**Fig 3.**
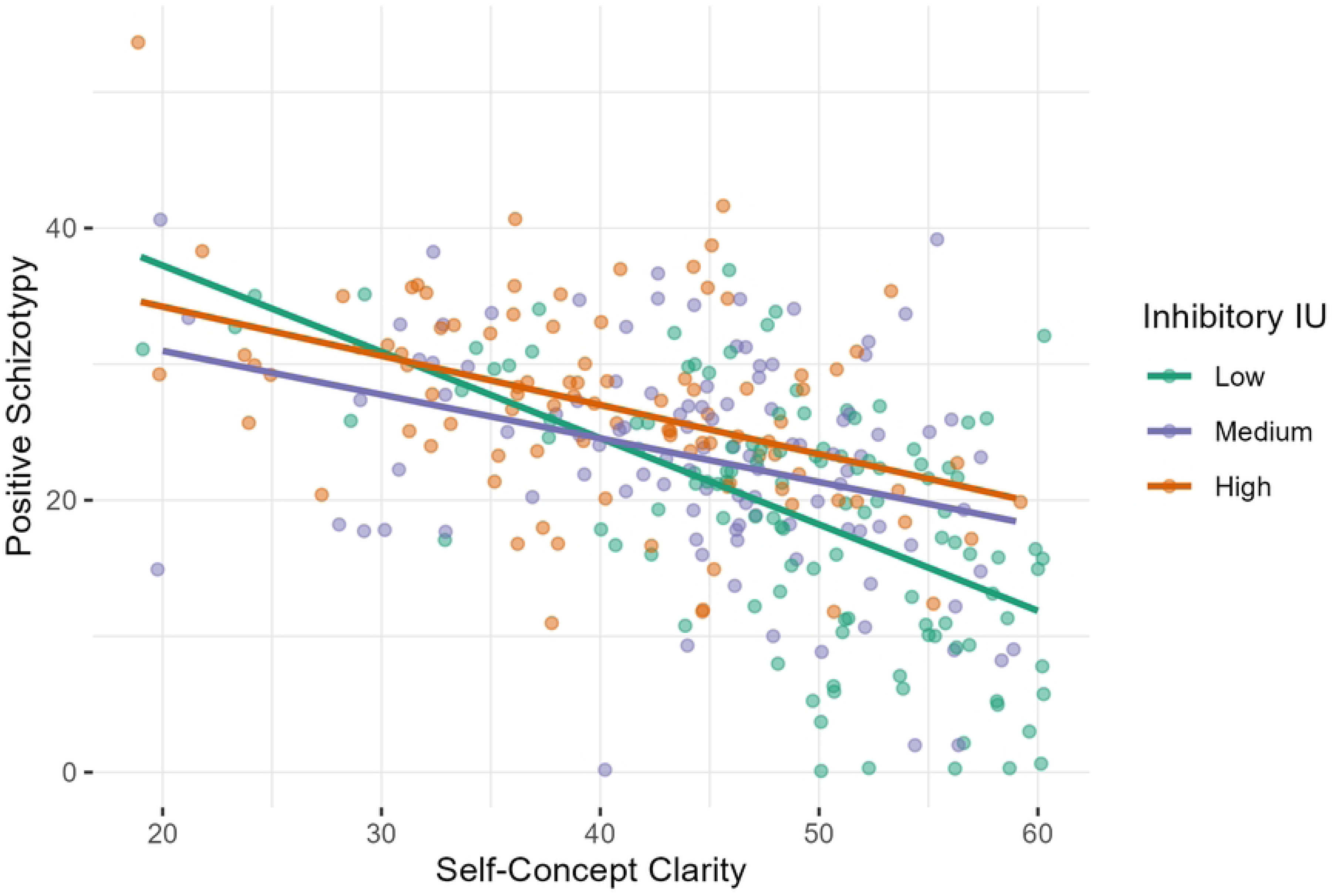
Interaction between SCCS and inhibitory IU predicting positive schizotypy. Note. SCCS = Self-Concept Clarity Scale. IU = Intolerance of uncertainty. Shaded lines represent low, medium, and high levels of IUS (based on tertile splits) with age, sex, and disorganized schizotypy as covariates.

Regarding the control variables, age and sex did not exhibit a significant effect in either model. However, the disorganized dimension demonstrated a significant influence (**prospective model**: *Beta = 0.999, SE = 0.066, Standardized Beta = 0.635, t = 15.162, p < 0.001*; **inhibitory model**: *Beta = 1.001, SE = 0.068, Standardized Beta = 0.637, t = 14.696, p < 0.001*). This was expected, as higher levels of disorganization may predict increased positive and negative traits, and vice versa [6]. However, the interaction remained significant even when controlling for this variable, indicating that the results cannot be explained solely by the central role of disorganisation.

## Discussion

Our findings provide valuable insights into the relationship between self-concept clarity, schizotypy, and intolerance of uncertainty. The negative associations between self-concept clarity and all three dimensions of schizotypy (positive, negative, and disorganized) suggest that individuals with lower self-concept clarity tend to exhibit higher schizotypal traits. This aligns with previous research indicating that disturbances in self-concept are characteristic to schizotypy and related psychopathological conditions [12,14–16]. The moderate strength of these associations further supports the notion that self-concept clarity is an important factor in understanding schizotypal traits.

The observed negative relationship between self-concept clarity and inhibitory intolerance of uncertainty suggests that individuals with a less coherent and stable self-concept may be particularly prone to experiencing difficulties in inhibiting actions or responses when faced with uncertainty. This finding is in line with theories suggesting that uncertainty can be particularly distressing for individuals with an unstable self-concept, leading to increased cognitive and emotional difficulties [19,24].

Our moderation analyses further highlight the complex interplay between self-concept clarity, intolerance of uncertainty, and positive schizotypy. The significant interactions suggest that inhibitory intolerance of uncertainty modifies the strength of the association between self-concept clarity and positive schizotypy. Specifically, individuals with lower levels of inhibitory intolerance of uncertainty exhibit a stronger negative relationship between self-concept clarity and positive schizotypy. In contrast, when inhibitory intolerance of uncertainty is high, this association becomes weaker. This finding suggests that low inhibitory intolerance of uncertainty may allow self-concept clarity to play a more decisive role in shaping positive schizotypal traits, potentially due to greater behavioral flexibility in the face of uncertainty. Conversely, individuals with high inhibitory IU may experience elevated levels of positive schizotypy regardless of self-concept clarity, possibly due to rigid avoidance tendencies and emotional overcontrol. These results underscore the importance of considering both self-structure and uncertainty tolerance in models of schizotypy and psychosis risk.

From a theoretical perspective, these findings support models that emphasize the role of self-concept disturbances in schizotypal traits. If we assume that reduced self-concept clarity causally triggers the emergence of positive schizotypy as a compensatory mechanism [27], our results suggest that tolerance of uncertainty may be a condition that facilitates the influence of self-concept clarity on positive schizotypy. Given that intolerance of uncertainty is a modifiable cognitive factor [39], these results have potential implications for interventions aimed at enhancing self-concept clarity and reducing the distress associated with uncertainty in individuals exhibiting schizotypal traits.

## Strengths and limitations

The present study offers several novel contributions to the understanding of schizotypal traits and their cognitive-affective underpinnings. First, it is among the first to examine whether intolerance of uncertainty moderates the relationship between self-concept clarity and positive schizotypy—an interaction that has not yet been explored in prior research. While both intolerance of uncertainty and self-concept clarity have been individually associated with psychopathology, particularly within anxiety and psychosis-spectrum disorders, their combined effect on schizotypal traits remains poorly understood. Second, by focusing on a non-clinical sample, this study contributes to the growing body of research supporting a dimensional view of psychosis and highlights how subclinical traits can be shaped by the interplay between identity-related and uncertainty-related cognitive processes. Finally, by integrating constructs typically studied in separate fields—such as self-concept clarity in identity development and intolerance of uncertainty in anxiety research—this study offers a more comprehensive framework for understanding vulnerability mechanisms that may precede the onset of clinical symptoms.

Although we aimed to gather a representative sample, demographic distributions suggest room for improvement. Notably, our sample includes over three and a half times more women than men, which does not reflect the general population. While previous research has shown that women are generally more willing to participate in studies [40], this imbalance may also be influenced by Facebook’s advertising algorithm. Since the algorithm’s mechanism is not fully transparent, we cannot determine the extent to which it distorted sample representation, whether it restricted visibility to certain groups, or how much it contributed to representativeness. Nonetheless, under current technical conditions, the use of Facebook advertising significantly increased participant recruitment and yielded a more heterogeneous sample in terms of age, education, and socioeconomic status compared to university-based recruitment.

Furthermore, while we conceptualized schizotypy as an analog for schizophrenia, our study did not include individuals with a clinical diagnosis. As a result, our findings cannot be directly generalized to clinical populations. However, a substantial body of research supports the notion that schizotypy reflects a continuum of liability for schizophrenia, both in terms of phenomenology and underlying neurocognitive mechanisms [2,41,42]. From this perspective, studying schizotypal traits in non-clinical populations offers a valuable window into the early and subclinical processes that may contribute to the development of psychotic disorders. We therefore believe that our findings provide a meaningful foundation for future research on schizophrenia and related conditions.

## Conclusions and future directions

We believe our findings have practical implications. Future research should explore these associations longitudinally to establish causal pathways and develop interventions aimed at enhancing self-concept clarity to mitigate schizotypal symptoms. Given that intolerance of uncertainty emerged as a moderating factor, targeted interventions could focus on reducing intolerance of uncertainty and fostering resilience in individuals at risk for schizophrenia. Improving one’s ability to manage anxiety related to uncertainty may, in turn, facilitate addressing self-related deficits that contribute to positive schizotypy.

In future studies, we aim to extend our research to clinical populations and compare clinical groups with subclinical individuals. Additionally, we plan to investigate the relationship between intolerance of uncertainty, positive schizotypy, and conspiracy beliefs, a highly relevant topic in contemporary psychological research.

## Data Availability

The dataset is accessible via the Open Science Framework (OSF) at the following link: https://osf.io/xafkn/?view_only=589c8c479f9447dc8b7de0c1085ddfab.

## Acknowledgements

MR, ÁS and DB were supported by the NAP2022-I-2/2022 Research Grant (Hungarian Brain Research Program 3.0). BP and ÁS were supported by the Janos Bolyai Research Scholarship of the Hungarian Academy of Sciences. Our data can be accessed through the following link: https://osf.io/xafkn/?view_only=589c8c479f9447dc8b7de0c1085ddfab

## Contributions

Conceptualization: AK, DB, ÁS, ÁV, MR, BP Data Curation: AK, DB

Formal Analysis: AK, BP Funding Acquisition: ÁS, MR, BP Investigation: AK

Methodology: AK, DB, ÁS, ÁV, MR, BP Project Administration: SK, BP Resources: DB, MR, BP

Software: AK, DB Supervision: SK, MR, BP Validation: BP Visualization: AK, BP

Writing – Original Draft Preparation: AK

Writing – Review & Editing: SK, DB, ÁS, ÁV, MR, BP

